# Human movement and transmission dynamics early in Ebola outbreaks

**DOI:** 10.1101/2023.12.18.23300175

**Authors:** Alexandria Gonzalez, Behnam Nikparvar, M. Jeremiah Matson, Stephanie N. Seifert, Heather D. Ross, Vincent Munster, Nita Bharti

**Author notes:** All authors declare no competing interests.

## Abstract

Human movement drives the transmission and spread of communicable pathogens. It is especially influential for emerging pathogens when population immunity is low and spillover events are rare. We digitized serial printed maps to measure transportation networks (roads and rivers) in Central and West Africa as proxies for population mobility to assess relationships between movement and Ebola transmission. We find that the lengths of roads and rivers in close proximity to spillover sites at or near the time of spillover events are significantly correlated with the number of EVD cases, particularly in the first 100 days of each outbreak. Early management and containment efforts along transportation networks may be beneficial in mitigation during the early days of transmission and spatial spread for Ebola outbreaks.

**Significance Statement:** This study links human movement and pathogen transmission across fifty years. While this relationship is well understood for modern outbreaks, it has not been characterized at local scales for historical outbreaks. We compared the number of cases and the spatial spread of each documented outbreak of Ebola (*Orthoebolavirus zairense*, EBOV) to the road and river networks surrounding each spillover location at the time of each spillover event. We measured the road and river networks by digitizing a series of paper maps that were printed during or near the year of each spillover. We show that the connectivity of spillover locations is consistently correlated to the severity of the outbreak over time and across all locations of EBOV spillover events.

## Introduction

Understanding the relationship between human mobility and the spread of pathogens is crucial for mitigating outbreak events. This has been demonstrated for acute respiratory infections, such as influenza and daily commuter patterns ^[1]^, for measles and large-scale seasonal movements ^[2]^, and for HIV with long term changes in connectivity ^[3]^. Understanding this relationship can highlight spatial and temporal risks of transmission, which can inform public health preparedness, surveillance, and outbreak response efforts.

The zoonotic origins and high case fatality rate in humans (40-90%) of Ebola virus (EBOV, member of species *Orthoebolavirus zairense,* formerly known as Zaire ebolavirus) are particularly concerning as a disease-causing agent ^[4,5]^. Possible reservoir hosts include fruit bats while non-human primates and some antelope are known intermediate hosts ^[4]^. Initial symptoms of Ebola virus disease (EVD) include headaches, high fever, and muscle aches. Disease progression can lead to more severe symptoms, such as internal bleeding and organ failure, which can result in death ^[4,6]^.

The first documented case of EVD caused by the EBOV occurred in Yambuku, DRC, 1976. From 1976-2020, a total of 18 human EVD spillovers were documented, all within Central and West Africa ^[7]^. A spillover is defined as a cross-species transmission event and often refers to pathogen transmission from wildlife to humans. Many EVD outbreaks are triggered by a single spillover event followed by human-to-human transmission between close contacts. Most spillover events have been traced to rural and heavily forested areas, where human-wildlife interactions could have occurred. Index cases are often linked to hunting activities, or individuals who work in forests or areas of land conversion (e.g., logging, mining, etc.) and may have had contact with bats or non-human primates ^[8,9]^. Infected humans can transmit the virus directly to other humans with the greatest risk to close contacts, including caregivers, who may be family members or medical professionals ^[10]^.

To identify possible links between human movement and the spread of EBOV, we measured movement and transportation networks across time and compared them to concurrent EVD outbreaks. Specifically, we analyzed characteristics of the road and river networks surrounding the spillover locations near the time of each spillover event for all 18 documented EVD outbreaks from 1976-2020. We quantified the connectivity of spillover locations to surrounding areas. Roads vary by condition and surface (e.g., unpaved roads) and enable transportation by vehicles, animals, and on foot ^[11,12]^. Rivers are also commonly used for transit, particularly in Central Africa, and link large cities as well as smaller settlements ^[12]^.

We first examined metrics for total outbreaks and next focused on the first 100 days of each outbreak to isolate the immediate importance of movement and connectivity. This temporal scope helped remove the downstream effects of transmission and long-term outbreak management. It also allowed us to isolate the early trajectories of two recent EVD outbreaks that were uncharacteristically large in number of cases, spatial extent, and total duration ^[10]^.

We also examined proxies for the impact of outbreak response readiness and prior experience by comparing the total outbreak size and duration for the first EBOV outbreaks in an area to subsequent outbreaks in the same location. Using the locations of index cases, we defined ‘subsequent outbreaks’ as EBOV spillover events that occurred in close spatial proximity to a previous spillover event, either in the same town or a neighboring town with prior evidence of connectivity due to movement (< 60 km). We identified three sets (two pairs and one triad) of first and subsequent outbreaks and compared their outbreak metrics.

We found that measures of transportation networks were correlated with several metrics used to measure total outbreaks. Measures of transportation networks included total road length, total combined road and river length, and total number of intersections (road-road, road-river, and river-river intersections). However, this relationship was even stronger for the first 100 days of outbreaks; transportation networks surrounding each EBOV spillover event were significantly positively correlated with the number of reported cases in the first 100 days of an outbreak. This suggests that population mobility is a contributing factor in EBOV transmission. We also found that subsequent outbreaks were always smaller than initial outbreaks, suggesting preparedness or response experience may improve outbreak management. With increasing human mobility and population connectivity, emerging zoonoses present a growing threat to populations and global health systems.

## Methods

### Data sources

We scanned and digitized printed versions of Michelin Road maps of Central and South Africa that were published in 1963, 1969, 1974, 1981, 1989, 2003, 2007, and 2019 and West Africa from 1975, 1989, 1991, 2003, and 2019 (**Table 1**) ^[13,14]^. We used shapefiles of administrative areas from *DIVA-GIS* as reference maps for each country ^[15]^. We included all countries with documented Ebola-Zaire virus spillovers or transmission events from 1976 to 2020: Central African Republic, The Democratic Republic of the Congo (DRC), Gabon, Guinea, Liberia, Republic of the Congo (RC), Sierra Leone, and Uganda (Fig. 1a). We used ArcMap *version 10.8.1* to georeference and digitize roads and rivers from each scanned map ^[16]^. Maps were georeferenced using the WGS 1984 geographic coordinate system and projected to the Africa Sinusoidal projected coordinate system. Serial road and river networks from different years were registered together to minimize the errors associated with digitizing. Topology corrections were applied on the networks in GRASS GIS *version 7.8.7* ^[17]^.

**Figure 1.**
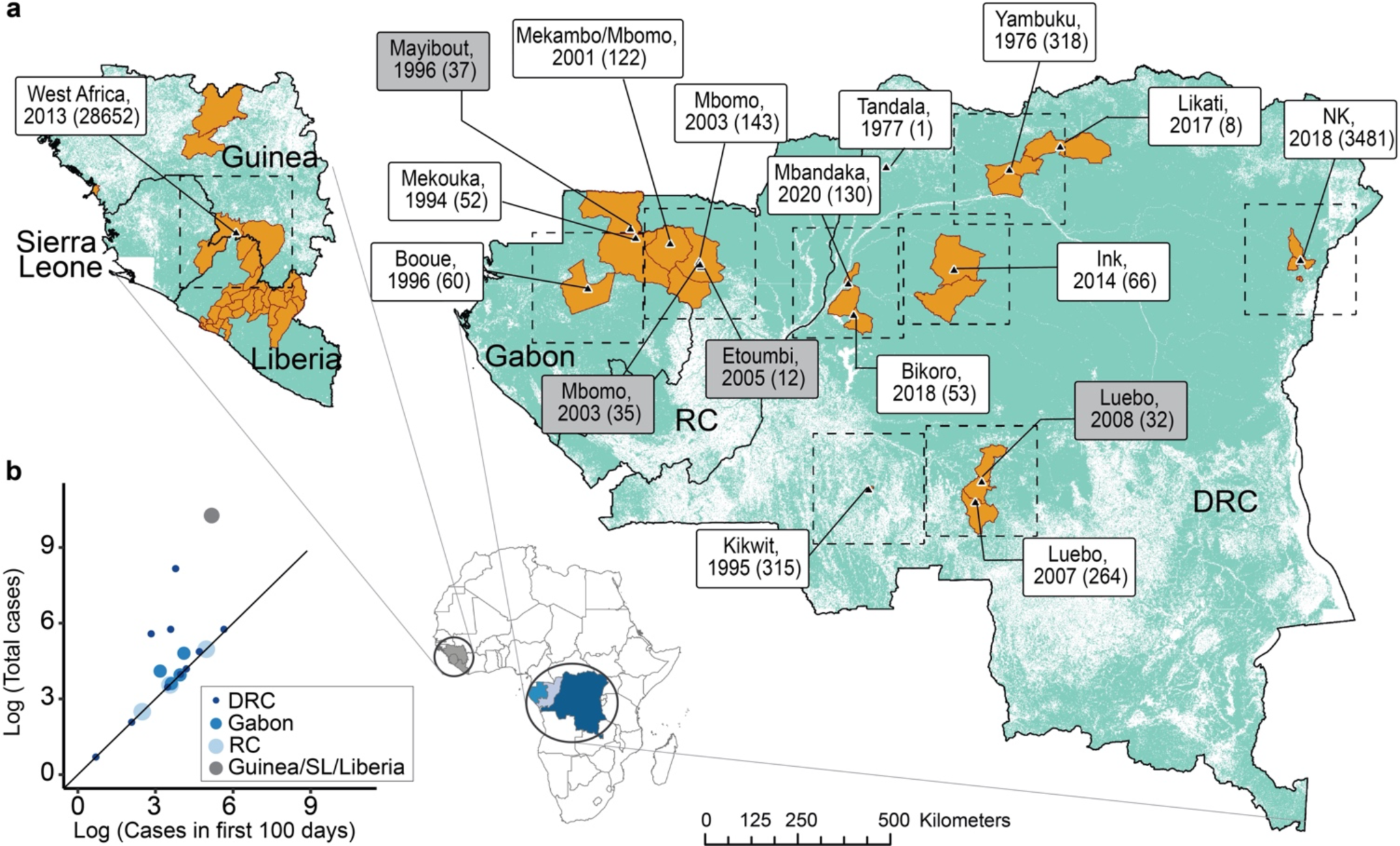
Spillover Events. **(a)** Locations of EBOV spillovers. Black triangles indicate locations of spillover events to humans, labeled with the location, year, and total cases in parentheses. Labels for subsequent outbreaks are shaded in gray. Orange filled polygons represent the estimated geospatial spread of each outbreak in the first 100 days of the outbreak; the West African outbreak shows the only non-contiguous area. **(b)** Outbreak measures: The total cases and the total cases in the first 100 days of each outbreak.

**Table 1.**
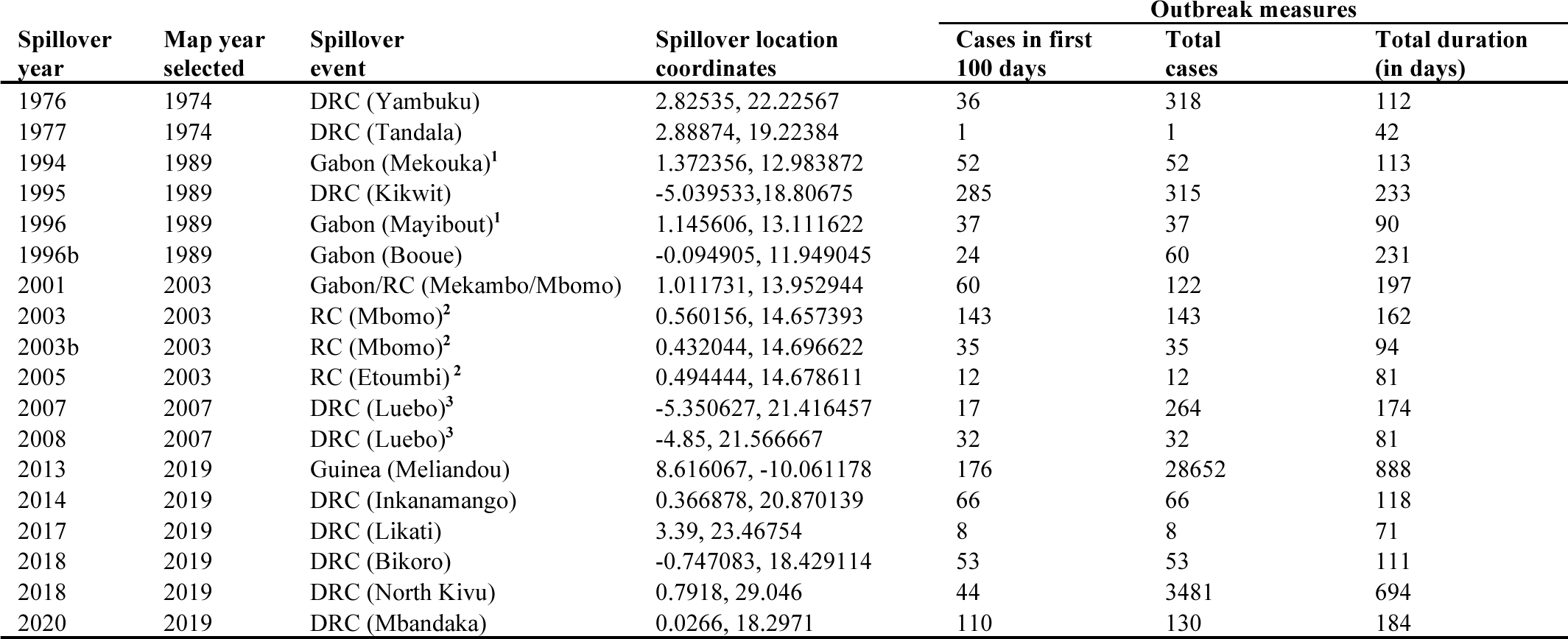
Summary of EBOV spillover events from 1976-2020 including the year, location, number of cases in the first 100 days, total cases, and total duration of each spillover. The map year selected represents the publication year of the Michelin map that was temporally closest to each spillover event. The three sets (two pairs and one triad) of initial and subsequent outbreaks are indicated with matching numerical superscripts.

We analyzed documented spillover events from the DRC that occurred in 1976, 1977, 1995, 2007, 2008, 2014, 2017, 2018, 2019, and 2020, Gabon in 1994, 1996, 1996(b), and 2001, RC in 2003, 2003(b), and 2005, and Guinea in 2013. We used the map year closest to each spillover event year to perform a detailed analysis on the road and river networks. The greatest time difference between a map year and the corresponding spillover year for an event was 7 years, and the average was 3 years. Table 1 includes spillover event details and the map year that was matched to each spillover event. We excluded outbreaks that were determined by genetic sequence analysis that were caused by transmission from a survivor of an outbreak in years prior; we included only outbreaks that were seeded by spillover events. We also excluded other orthoebolaviruses, which differ in incubation period, transmissibility, clinical presentation, and geographic range.

### Data preparation

We measured outbreaks using the total number of cases recorded during the outbreak, the duration of each outbreak in days, and the number of cases in the first 100 days (Fig. 1b) for Yambuku ^[18,19]^, Tandala, Mekouka ^[20]^, Kikwit ^[20,21]^, Mayibout ^[20]^, Booue ^[20,22]^, Mekambo/Mbomo ^[23,24]^, Mbomo ^[25,26]^, Etoumbi ^[27]^, Luebo ^[28,29]^, Meliandou ^[30,31]^, Inkanamango ^[32]^, Likati ^[33,34]^, Bikoro ^[35,36]^, North Kivu ^[37,38]^, and Mbandaka ^[39]^. We also examined the spatial spread of notified cases within the first 100 days of each outbreak from existing maps and surveillance reports in above cited references.

We performed a sensitivity analysis of correlation between transportation networks and outbreak measures across study areas with each spillover event at the center. We examined square study areas of 50 km x 50 km, 100 km x 100 km, 150 km x 150 km, 200 km x 200 km, and 300 km x 300 km. We assessed correlations between measures of mobility networks and outbreaks at 10 km increments for sensitivity analysis (Fig. S1).

We used estimates of walking distance and speed to determine the study area that would capture movement during the shortest incubation period for EVD. We apply the operational incubation period for EVD during outbreak response, which is 2-to-21-days, although 4-5 day minimum incubation periods are more typical than 2 days ^[40,41]^. An average person is able to walk 5 km/hr along roads ^[42]^. The estimated travel speed by boat is in the same range as walking speed ^[10]^. Thus, at 7.5 hours of walking or traveling by boat per day, a person could potentially travel 75 km in 2 days. We concluded that a study area of 75 km around each spillover, or a 150 km x 150 km area centering each spillover, would adequately represent the minimum critical movement early in an epidemic and we focus our results accordingly (see SI for results from remaining study areas). We calculated the total length of the roads and rivers within each study area. Additionally, we quantified the number of total intersections for each study area where an intersection is defined as a point where at least two routes intersect (e.g road-road, road-river, river-river).

### Subsequent Outbreaks

We defined subsequent outbreaks as EBOV spillover events that occurred in the same location or within close spatial proximity, no greater than 60 km apart, from a previous spillover event. These locations also exhibited evidence of connectivity, including pathogen transmission between them and direct links along transportation networks ^[10,43]^. We identified three sets of subsequent outbreaks. First, an outbreak in Mekouka, Gabon in December 1994 preceded an outbreak in Mayibout, Gabon in February of 1996. Second, an outbreak in Mbomo, RC in February 2003 preceded both outbreaks in Mbomo in November 2003 and Etoumbi, RC in May 2005. Third, an outbreak in Luebo, DRC in September 2007 preceded an outbreak in Luebo in December 2008 (Fig. 1a).

### Statistical Analysis

We calculated Pearson correlation values to assess the relationships between outbreak measures and transportation network characteristics. We used linear regressions and multiple regression models to assess the relationship between each outbreak measure and the transportation network characteristics described. We also added the total road networks and the total river networks separately to a multiple regression analysis against the total cases in the first 100 days of each outbreak for the 150 km x 150 km study areas. We reported R-squared and p-values for each analysis. We used R studio, version 2022.07.1+554 for all statistical analyses ^[44]^.

## Results

We mapped each EBOV spillover event (Fig. 1a). Between 1976 and 2020, 10 EBOV spillover events occurred in the DRC, 3 in the ROC, 4 in Gabon, and 1 in Guinea. The total number of cases for each outbreak ranged from 1 to 28,652 (Table 1) ^[5,7]^. The Guinea outbreak of 2013 reported 28,652 cases and a duration of 888 days; this outbreak represents the largest number of total cases and the longest duration of any EVD outbreak, while the Tandala outbreak in 1977 reported the fewest total cases (1 case) and the shortest duration (42 days). We also mapped the corresponding estimated geospatial spread for the first 100 days of each outbreak ^[43]^. The total cases in the first 100 days of each outbreak ranged from 1 to 285; the Kikwit outbreak of 1995 reported the most cases in the first 100 days with 285 cases.

### Network characteristics

We measured the lengths of the road and river networks around each reported spillover location from the Central/South Africa and North/West Africa Michelin maps from 1963-2019 and 1975-2019, respectively. Across the 150 km x 150 km study areas in Central Africa, from 1963 to 2019, we observed a maximum increase in road network length of 363 km and a maximum decrease in road network length of 120 km for 17 spillover events. The average road network length change was an increase of 53 km (see Table S1 for the net changes in road lengths in a 150 km x 150 km study area surrounding each spillover). The change in road network length from 1975 to 2019 in the 150 km x 150 km study area surrounding the West Africa spillover event was an increase of 133 km. River lengths were largely stable over time within each study area.

We calculated the ratio of river network length to road network length in a 150 km x 150 km study area surrounding each spillover (Fig. 2). The study area surrounding the 1994 outbreak in Mekouka, Gabon contained the highest ratio of river to road length, at 853 km of rivers to 61 km of roads, while the study area around the 1977 spillover event in Tandala contained the lowest ratio of rivers to roads, at 260 km of rivers to 710 km of roads. The Kikwit outbreak event of 1995 had the most road length with 1196 km. There was no temporal relationship between the year of spillovers and the ratio of river to road network length. At larger spatial scales, such as 300 km x 300 km, the ratio of river length to road length was approximately 1:1 for most outbreaks.

**Figure 2.**
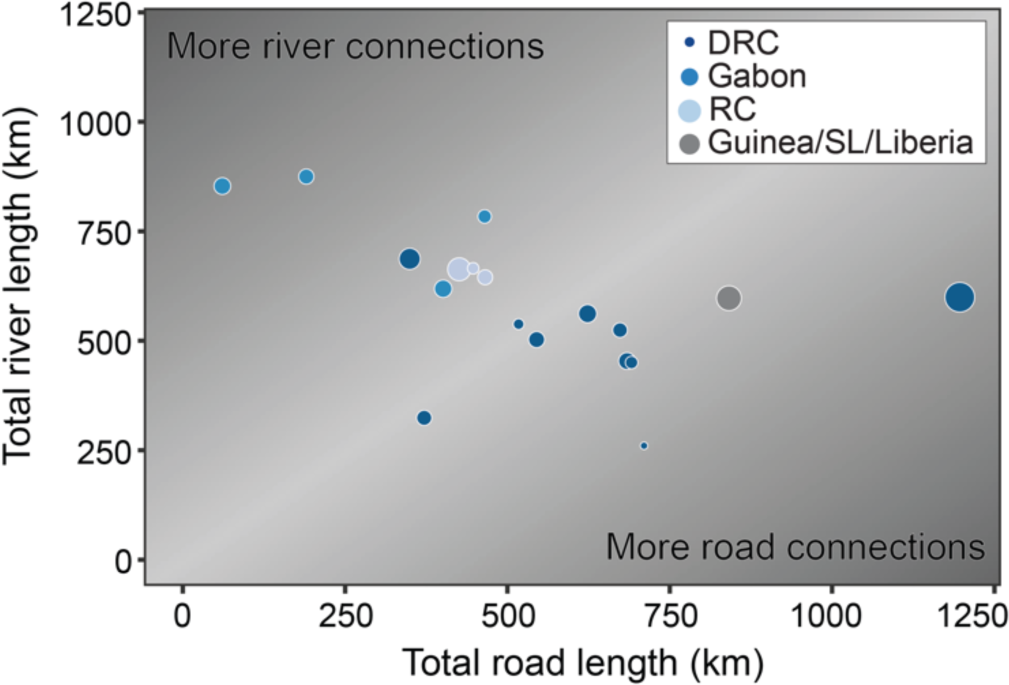
Total road and river network lengths for each spillover event at the time of the spillover event in a 150×150 study area surrounding the spillover. The 1:1 line is shaded in the lightest gray in the background, size of points corresponds to number of cases in the first 100 days of each outbreak, and color corresponds to location of spillover.

### Total cases and outbreak duration

We examined the relationship between the transportation networks and outbreak metrics, specifically the total number of cases and total duration in days for each outbreak, using linear regression models. We conducted two separate analyses: one including all spillover events and one excluding the large Guinea and North Kivu spillover events.

#### All spillover events

In 150 km x 150 km study areas, the R-squared values indicated that total river network lengths, total road network lengths, total combined road and river network lengths, and total number of intersections each explained between 0 - 12% (p > 0.05) of the variation in the total cases and total duration in days of the outbreaks. Within the 100 km x 100 km study area, the R-squared values indicated that the total combined road and river network lengths explained 28% (p = 0.0236) of the variation in total cases and 35% (p = 0.0101) of the variation in total duration in days (Fig. S2). Within the 300 km x 300 km study area, the R-squared values indicated that the total combined road and river network lengths explained 23% (p = 0.0434) of the variation in total cases. We found no significant relationships between transportation networks and outbreak metrics for the remaining study areas (see Table S2 for complete results).

#### Excluding Guinea and North Kivu spillover events

We found that the two largest and longest EBOV, the Guinea (2013; 28652 cases in 888 days) and North Kivu (2018; 3481 cases in 694 days) events (Table 1), occasionally obscured the relationship between transportation networks and both the total cases and total duration in days. When excluding these outbreaks in 150 km x 150 km study areas, the R-squared values indicated that total river network lengths, total road network lengths, total combined road and river network lengths, and total number of intersections each explained between 4 – 25% (p > 0.05) of the variation in the total cases and total duration in days of the outbreaks. Within the 200 km x 200 km study areas, the R-squared values indicated that the total combined road and river network lengths and the total road network lengths each explained 29% (p = 0.0320) of the variation in the total cases. Additionally, the total combined road and river network lengths explained 28% (p = 0.0354) of the variation in outbreak duration within this study area. When excluding the two largest outbreaks in our analyses of 300 km x 300 km study areas, the R-squared values indicated that the total number of intersections explained 27% (p = 0.0380) of the variation in the total cases and the total river length explained 57% (p = 0.0007) of the variation in the total duration. See Table S3 for complete R-squared values.

### The first 100 days of each outbreak

We examined the relationship between the transportation networks and the total cases in the first 100 days of each outbreak (Table 1 and Table S2-5). The total combined road and river network lengths were strongly correlated with the total number of intersections for all of the study areas examined: 50 km x 50 km, 100 km x 100 km, 150 km x150 km, 200 km x 200 km, and 300 km x 300 km (r = 0.81 – 0.94). We found that the total combined road and river network lengths, explained between 35 – 56% of the variance of the total cases in the first 100 days across all outbreaks for each of the study areas examined: 50 km x 50 km, 100 km x 100 km, 150 km x150 km, 200 km x 200 km, and 300 km x 300 km (Table S2). We observed similar results with the number of intersections and the total cases in the first 100 days for each of the study areas. The total road network lengths explained between 27 - 36% of the variance of the total cases in the first 100 days of an outbreak for the following study areas: 150 km x 150 km, 200 km x 200 km, and 300 km x 300 km. The total river network lengths explained between 2 - 22% (p > 0.05) of the variance of the total cases in the first 100 days of an outbreak for each of the study areas examined. The strongest relationship was between the total combined road and river network lengths and the total cases in the first 100 days for each of the study areas examined.

In 150 km x 150 km study areas, the R-squared values indicated that the total combined road and river network lengths and total road length explained 54% (p = 0.0005) and 30% (p = 0.0183) of the variation in the total cases in the first 100 days of each outbreak, respectively (Fig. 3). The total number of intersections against the total cases in the first 100 days is shown in Fig. S3. The total river network length in this study area explained less than 3% of the variation in the number of cases during the first 100 days of each outbreak. See Table S2 for complete R-squared values.

**Figure 3.**
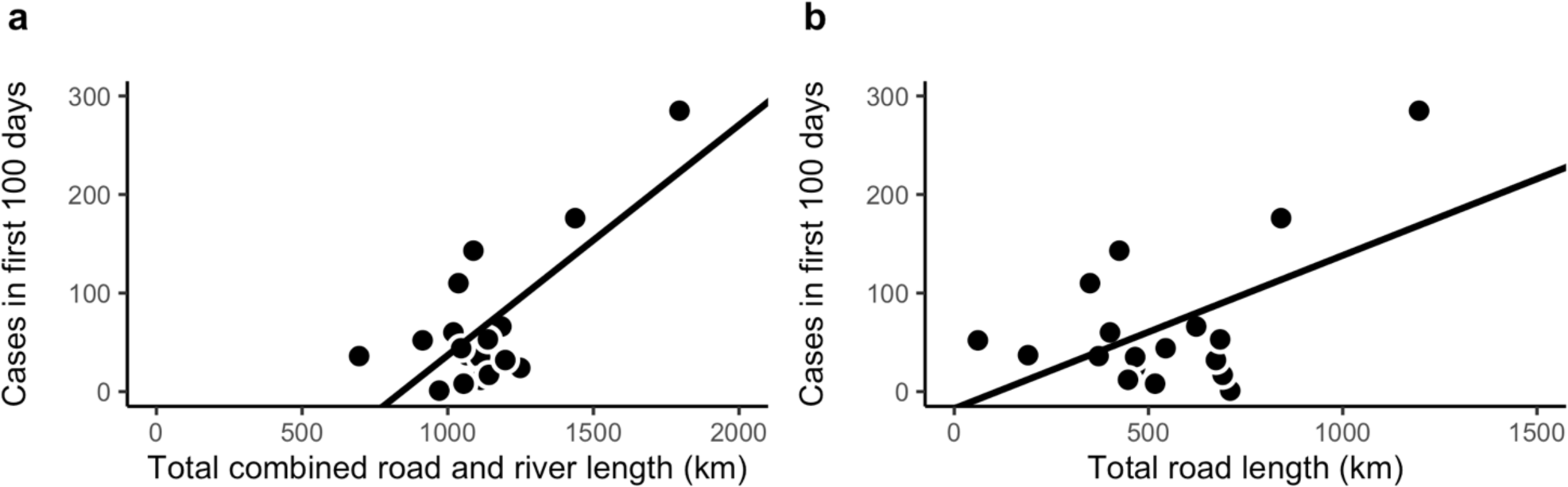
Linear Regression model on 150 km x150 km study area. **(a)** Total cases in the first 100 days of each outbreak against the total combined road and river length in km (R-squared = 0.5377, p = 0.0005) **(b)** Total cases in the first 100 days of each outbreak against the total road length in km (R-squared = 0.3012, p = 0.0183).

### Multiple regression analysis

We found that the total road network lengths and the total river network lengths added separately to a multiple regression model explained 54% (p = 0.0030) of the variation in the number of cases during the first 100 days of each outbreak. These results are similar to those that we observed when we analyzed the relationship between total combined road and river network lengths and total cases in the first 100 days using a linear regression model, as described above. The R-squared values also indicated that the total road network length alone explained 30% (p = 0.0183) of the variation in the total cases in the first 100 days of each outbreak.

Complete R-squared values for the transportation networks against outbreak measures including total cases, total duration in days, and total cases in the first 100 days at each study area is found in Table S2. Additionally, Fig. S4a-d shows the total combined road and river network lengths against the total cases in the first 100 days for the following study areas: 50 km x 50 km, 100 km x100 km, 200 km x 200 km, and 300 km x 300 km.

### Subsequent outbreaks

Each of the 4 outbreaks that were classified as ‘subsequent outbreaks’ reported fewer total cases and a shorter duration in days than the preceding outbreak that occurred in the same or similar location. Each subsequent outbreak occurred within 2 years and < 60 km of the first outbreak. The shortest elapsed time between an initial and subsequent spillover event was 268 days while the longest elapsed time was 471 days. The subsequent outbreaks reported between 15 and 232 fewer cases than their preceding outbreaks. The subsequent outbreaks were between 15 and 108 days shorter in total duration than their associated initial outbreaks. Following the initial Mbomo outbreak in February 2003, the subsequent outbreaks in Mbomo in November 2003 and Etoumbi in May 2005, respectively, reported approximately 75% and 90% fewer cases compared to the total cases reported in the first outbreak. Likewise, the total duration of these subsequent outbreaks decreased by over 40% and 50%, respectively, compared to the duration of the initial spillover event (Table 1, Table S4).

## Discussion

This study provides a novel data resource and novel analyses of transportation infrastructure surrounding all documented EBOV spillover events through 2020. All 18 of these events occurred within Central and West Africa. We examined the relationship between transportation infrastructure, which we used as a proxy for population mobility and connectivity, and outbreak measures. For outbreak measures, we considered the total number of cases in each outbreak, the total duration of each outbreak measured in days, and the number of cases reported in the first 100 days of each outbreak. We observed that the transportation networks had a stronger relationship with the number of cases in the first 100 days of an outbreak compared to the outbreak totals. Transportation network characteristics did not consistently show a significant relationship with the total cases or the total duration in days for each outbreak, likely because additional factors, including outbreak management, become determining factors in outbreak size after 100 days. These results suggest that transportation networks may play an important role in the transmission dynamics of EVD in the early stages of an outbreak.

Transportation networks connect people across locations. These networks both influence and are influenced by human mobility. Some EBOV-infected patients travel to healthcare centers and unknowingly transmit the virus along the way while others transmit the virus to healthcare personnel or visitors at the hospital ^[45]^ while others believe they are uninfected and travel to escape an outbreak. The geographic spread of EVD following a spillover event is determined by the hosts’ ability to travel. This is consistent with the outbreaks that reported the greatest numbers of cases (e.g., West Africa) and large geographic spread ^[46]^.

The influence of road networks of river networks on movement depends on various factors. Previous studies found that small villages are especially subject to poor road upkeep which limits the utilization of vehicles and hinders efficient travel ^[47]^. Individuals in these areas will instead use animals like donkeys or walking as means of travel along roads ^[11]^. River networks are also commonly used for transportation. Rivers connect major cities, for example Kinshasa and Kisangani in DRC, and Brazzaville and Kinshasa between DRC and RoC and also provide critical links between many smaller settlements^[10,12,48]^.

To examine the immediate importance of movement and connectivity as it relates to outbreak severity, we conducted a targeted analysis on the first 100 days of each outbreak. This approach provided insight into the short-term effects of transmission and early outbreak management rather than the downstream effects of prolonged exponential outbreak growth and long-term outbreak management. This approach highlighted the potential importance of early interventions in outbreak containment. In addition, the largest outbreaks (West Africa 2013 and North Kivu 2018) were no longer outliers when using this approach (Fig. 1b). This analysis allowed for a comprehensive examination of the early mechanisms that underlie each outbreak despite the sizable differences in the total number of cases and duration in days across outbreaks. Due to the very high proportion of susceptible individuals to EBOV infection in most populations, movement patterns early in an outbreak are extremely important in determining outbreak transmission dynamics. We find strong positive correlations between the transportation networks and EVD incidence in the first 100 days of each outbreak at various study areas. These findings provide insight into the relationship between human mobility and the spread of Ebola virus and further suggest that rapid response measures along transportation networks surrounding spillover events during the initial stages of an outbreak have the potential to reduce outbreak size.

Previous research identified an association between delayed recognition of EVD and longer, larger outbreaks. We observed similar results in the context of a small number of subsequent outbreaks which occurred in the ROC, Gabon, and the DRC (Fig. 1a). We found that even when spillover events occur in close spatial proximity and close together in time, and therefore rely on very similar or identical road and river networks, the subsequent outbreak resulted in less severe outbreaks in terms of total cases, total duration (in days), and spatial spread. This was most strongly demonstrated by the two subsequent spillover events in Mbomo in November 2003 and Etoumbi in May 2005 that followed the initial Mbomo outbreak in February 2003. With each of these subsequent spillover events, we observe fewer total cases and shorter duration, indicating a favorable trend towards efficient containment and control of the outbreaks. Excluding subsequent events from the regression analysis strengthened the positive association between transportation networks and both total cases and total outbreak duration for many scenarios (Table S2 and S4). The responses to subsequent spillover events, especially those that occur shortly after the first spillover, may have benefited from experiential knowledge and outbreak response infrastructure developments, such as improved surveillance and diagnostics. This suggests that preparedness and rapid response, including early outbreak recognition, maybe be able to overcome the impact of movement in outbreak trajectory. We performed an additional analysis between transportation networks and total cases and total outbreak duration in which we excluded both subsequent events and outlier events. We found that the positive association between transportation networks and total cases and total duration was often further increased when excluding both subsequent and outlier events (Table S2 and S5).

To control future outbreaks, management plans can benefit from the inclusion of human movement and population connectivity, particularly in the early stages of outbreaks. Land use change (e.g., deforestation due to logging, mining, and development) has drastically increased pressures on ecosystems in parts of Central and West Africa. One study found that the index cases in humans of EVD outbreaks (2004-2014) occurred mainly in forest fragmented hotspots ^[49]^. As population mobility and human settlements expand due to an increased demand for services and resources ^[50]^, forest fragmentation and wildlife disruption are an imminent concern with potentially deadly consequences.

In addition to enabling the geographic spread of pathogen transmission following a spillover event, movement may also assist in the occurrence of spillover events. Areas along rivers can have an abundance of fruit trees, which are common feeding sites for migrating fruit bats. For example, during the 2007 EBOV spillover event in Luebo, DRC, fruit bats suspected of being EBOV reservoirs (*H. monstrosus* and *E. franqueti*) migrated along the Lulua River near Luebo, stopping at settlements along the river, where residents handled them ^[51]^. The influence of transportation networks and population mobility on the emergence and transmission of EBOV and other transmissible pathogens is wide-reaching and would benefit from further exploration.

Control efforts, particularly in the early days of an outbreak are critical in infectious disease outbreak management, including EBOV. These efforts include strengthening infection prevention & control (IPC) programs (this may include testing and contact tracing along roads and rivers), strengthening disease surveillance systems, improving health infrastructure, and developing accessible public health communications ^[52]^. Additional local factors that must be considered in control efforts include political and economic instability and conflicts, displaced populations, and large-scale population movements ^[50]^. Experiential knowledge gained from managing spillover events of other zoonotic pathogens can also serve to help guide control efforts within the early stages of an outbreak in the face of future threats.

## Acknowledgments

We thank Kelsee Baranowski, Christina Faust, and Ephraim Hanks for their valuable feedback and insight in the development of this manuscript.

## Contributions

NB and VM were responsible for the concept of the study. NB, AG, and BN completed the analyses. VM, MJM, and SNS provided relevant insight, data, and feedback. HDR provided support for map discovery, archiving, and digitization. NB and AG prepared the first draft of the manuscript. All authors reviewed and approved the manuscript before submission.

## Funding

This study was supported by the joint National Institutes of Health (NIH) - National Science Foundation (NSF) - National Institute of Food and Agriculture (NIFA) Ecology and Evolution of Infectious Disease (award R01TW012434 to NB), NSF RAPID (award 2202872 to NB), and the Intramural Research Program of the National Institute of Allergy and Infectious Diseases (NIAID), NIH (1ZIAAI001179-01 to VM). SNS was partly supported by funding to Verena (viralemergence.org) from the NSF (award BII 2021909 and BII 2213854). Funders had no role in the study design, data collection and analysis, decision to publish, or preparation of the manuscript.

## Data availability

Hard copies of all Michelin maps used in this study are available at the Donald W. Hamer Center for Maps and Geospatial Information in the Penn State University Pattee Library. Digital copies and code used for analyses are available at https://github.com/bhartilab/EbolaMaps.

## Supplementary Information

**Figure S1.**
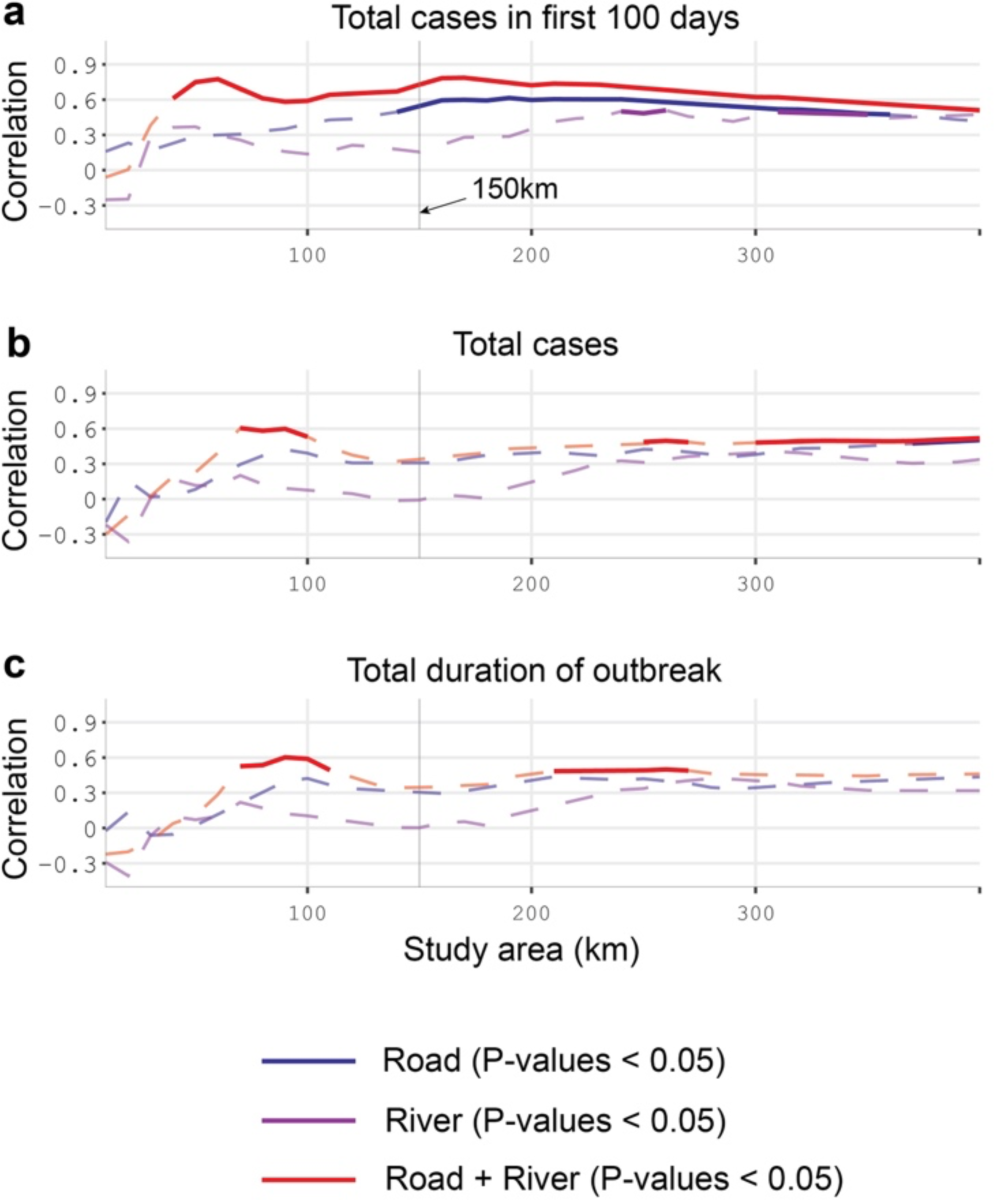
Sensitivity analysis of study area for correlations between transportation networks and outbreak measures: (A) cases in first 100 days, (B) total cases, and (C) total duration of outbreak. Dashed lines show p-values > 0.05; solid lines show p-values < 0.05; colors indicate transportation network.

**Table S1.** Net change in road length from 1963 to 2019 in the 150 km x 150 km study area surrounding the 17 spillover events that occurred in Central Africa. (+) Gain of road length, (-) loss of road length.

| Spillover location | Net change in road length (km) |
| --- | --- |
| Yambuku | + 362.99 |
| Tandala | + 105.67 |
| Mekouka | + 9.48 |
| Kikwit | + 184.77 |
| Mayibout | + 0.77 |
| Booue | + 52.49 |
| Mekambo/Mbomo | + 57.48 |
| Mbomo1 | + 13.78 |
| Mbomo2 | + 0.93 |
| Etoumbi | + 14.42 |
| Luebo07 | + 24.31 |
| Luebo08 | + 15.59 |
| Inkanamango | - 5.25 |
| Likati | - 37.86 |
| Bikoro | + 162.38 |
| North Kivu | - 119.55 |
| Mbandaka | + 13.83 |

**Table S2.**
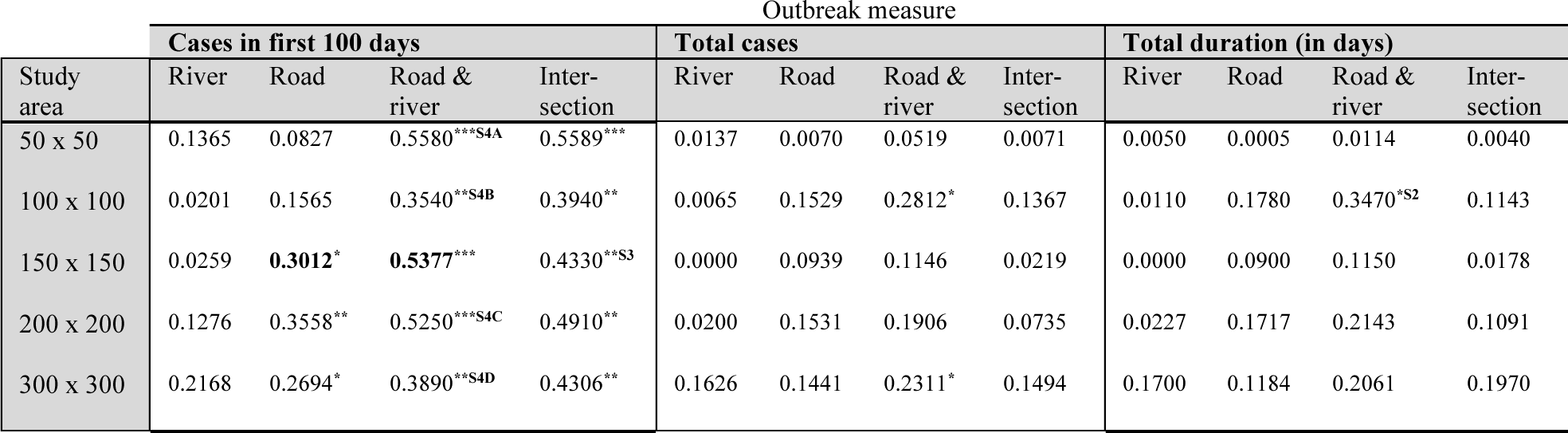
R-squared values for the transportation networks including total river length, total road length, total combined road and river length, and total intersections against outbreak measures: cases in first 100 days, total cases, and total duration in days for each study area examined (*p < 0.05, **p<0.01, ***p<0.001). All spillover events included. Upper case superscripts refer to figures that illustrate each relationship. Bold values correspond to Fig. 3A and Fig. 3B of the main paper, which illustrate these relationships.

**Table S3.**
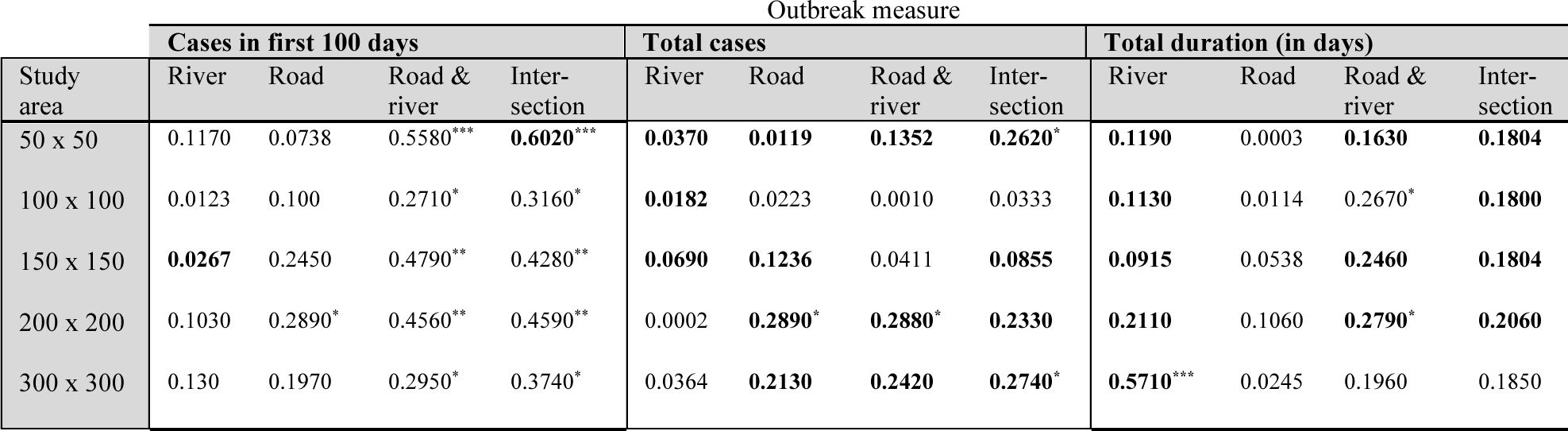
R-squared values for the transportation networks including total river length, total road length, total combined road and river length, and total intersections against outbreak measures: cases in first 100 days, total cases, and total duration in days for each study area examined (*p < 0.05, **p<0.01, ***p<0.001). Outlier events excluded. The outlier events are as follows: Meliandou Guinea in 2013 and North Kivu DRC in 2019. Upper case superscripts refer to figures that illustrate each relationship. Bold values are greater than when all spillover events are included.

**Table S4.** R-squared values for the transportation networks including total river length, total road length, total combined road and river length, and total intersections against outbreak measures: cases in first 100 days, total cases, and total duration in days for each study area examined (*p < 0.05, **p<0.01, ***p<0.001). Subsequent events excluded. The subsequent events are as follows: Mayibout Gabon in February1996, Mbomo RC in November 2003, Etoumbi RC in May 2005, and Luebo DRC in September 2007. Upper case superscripts refer to figures that illustrate each relationship. Bold values are greater than when all spillover events are included.

| Study area | Outbreak measure |  |  |  |  |  |  |  |  |  |  |  |
| --- | --- | --- | --- | --- | --- | --- | --- | --- | --- | --- | --- | --- |
|  | Cases in first 100 days |  |  |  | Total cases |  |  |  | Total duration (in days) |  |  |  |
|  | River | Road | Road & river | Inter-section | River | Road | Road & river | Inter-section | River | Road | Road & river | Inter-section |
| 50 x 50 | **0.1780** | **0.1340** | **0.6140\*\*\*** | **0.6110\*\*\*** | **0.0180** | **0.0090** | **0.0520** | **0.0073** | **0.0070** | 0.0004 | 0.0110 | **0.0043** |
| 100 x 100 | **0.0613** | 0.1560 | **0.3800\*** | 0.3720* | **0.0202** | **0.1620** | **0.2910\*** | 0.1250 | **0.0452** | 0.1760 | **0.3770\*** | 0.0916 |
| 150 x 150 | **0.0831** | **0.3160\*** | **0.5910\*\*** | 0.4300* | **0.0020** | 0.0920 | **0.1180** | 0.0162 | **0.0111** | 0.0760 | **0.1260** | 0.0085 |
| 200 x 200 | **0.2030** | 0.3540* | 0.5200** | 0.4650** | **0.0340** | 0.1490 | 0.1820 | 0.0600 | **0.0482** | 0.1470 | 0.1920 | 0.0760 |
| 300 x 300 | **0.2430** | **0.3260\*** | **0.4280\*** | **0.4490\*\*** | **0.1770** | **0.1620** | **0.2390** | 0.1470 | **0.1930** | **0.1400** | **0.2230** | 0.1930 |

**Table S5.** R-squared values for the transportation networks including total river length, total road length, total combined road and river length, and total intersections against outbreak measures: cases in first 100 days, total cases, and total duration in days for each study area examined (*p < 0.05, **p<0.01, ***p<0.001). Subsequent and outlier events excluded. The subsequent events are as follows: Mayibout Gabon in February1996, Mbomo RC in November 2003, Etoumbi RC in May 2005, and Luebo DRC in September 2007. The outlier events are as follows: Meliandou Guinea in 2013 and North Kivu DRC in 2019. Upper case superscripts refer to figures that illustrate each relationship. Bold values are greater than when all spillover events are included.

| Study area | Outbreak measure |  |  |  |  |  |  |  |  |  |  |  |
| --- | --- | --- | --- | --- | --- | --- | --- | --- | --- | --- | --- | --- |
|  | Cases in first 100 days |  |  |  | Total cases |  |  |  | Total duration (in days) |  |  |  |
|  | River | Road | Road & river | Inter-section | River | Road | Road & river | Inter-section | River | Road | Road & river | Inter-section |
| 50 x 50 | **0.1490** | **0.1210** | **0.6070\*\*** | **0.6460\*\*** | **0.0514** | **0.0203** | **0.1550** | **0.3090** | **0.1800** | **0.0006** | **0.1920** | **0.2210** |
| 100 x 100 | **0.0418** | 0.1070 | 0.3210 | 0.2970 | 0.0043 | 0.0124 | 0.0043 | 0.0145 | **0.3470\*** | 0.0033 | **0.3860\*** | **0.1590** |
| 150 x 150 | **0.0782** | 0.2620 | **0.5390\*** | 0.4200* | **0.0334** | **0.1150** | 0.0568 | **0.0607** | **0.2980** | 0.0399 | **0.3360\*** | **0.1640** |
| 200 x 200 | **0.1650** | 0.2940 | 0.4570* | 0.4370* | 0.0050 | **0.2800** | **0.2800** | **0.1870** | **0.4380\*** | 0.0776 | **0.2820** | **0.1620** |
| 300 x 300 | 0.1510 | 0.2580 | 0.3450* | 0.4050* | 0.0484 | **0.3150** | **0.3220** | **0.3100** | **0.7930\*\*\*** | 0.0460 | **0.2740** | **0.2150** |

**Figure S2.**
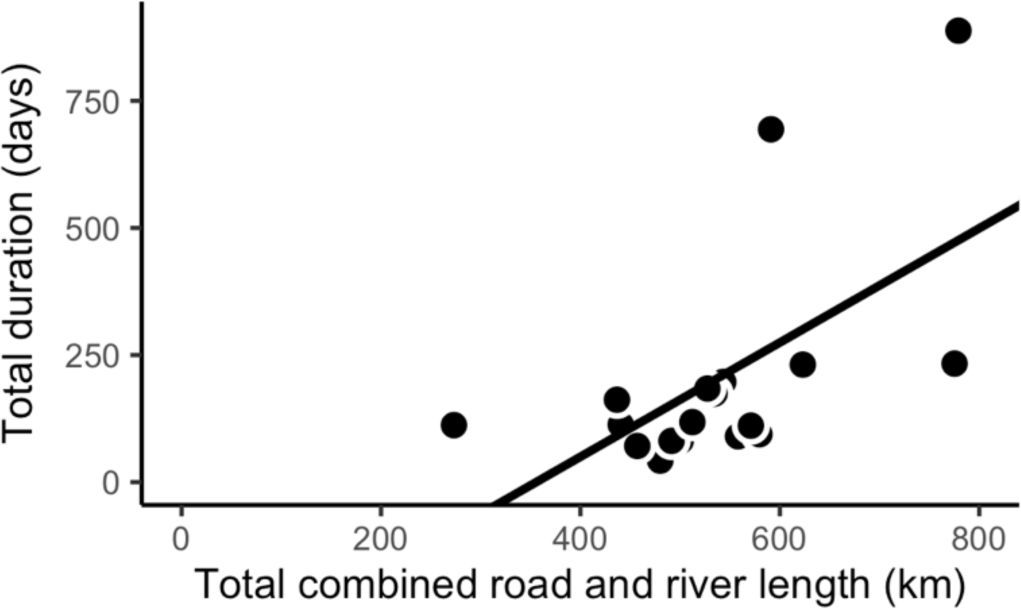
Linear Regression model of the total duration in days of each outbreak and the total combined road and river length in km in each 100×100 study area surrounding the spillovers (R-squared = 0.3470, p = 0.0101).

**Figure S3.**
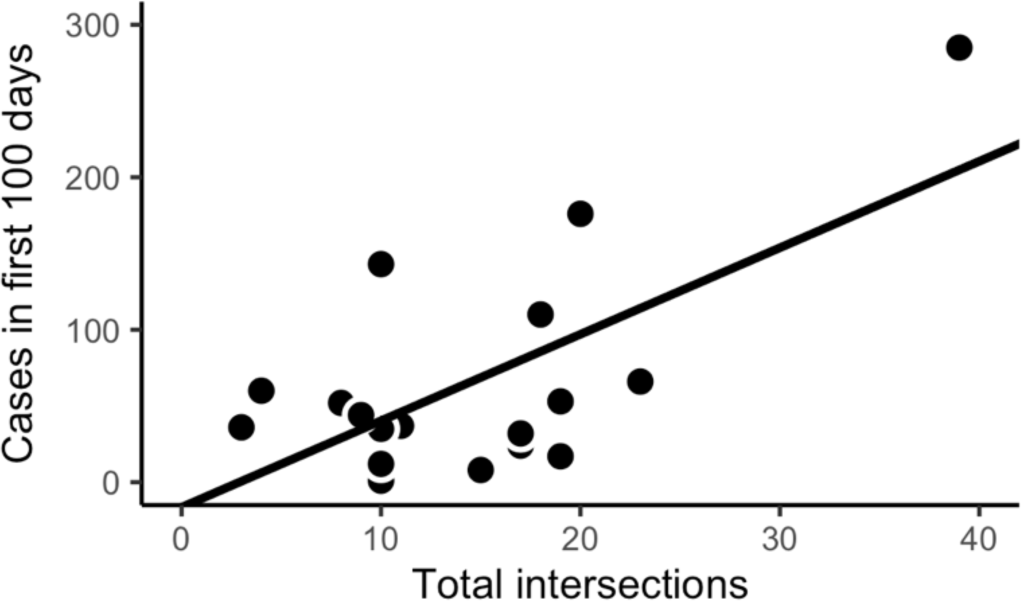
Linear Regression model of the total cases in the first 100 days of each outbreak and the total number of intersections in a 150 km x 150 km study area surrounding the spillovers (R-squared = 0.4338, p = 0.0030).

**Figure S4.**
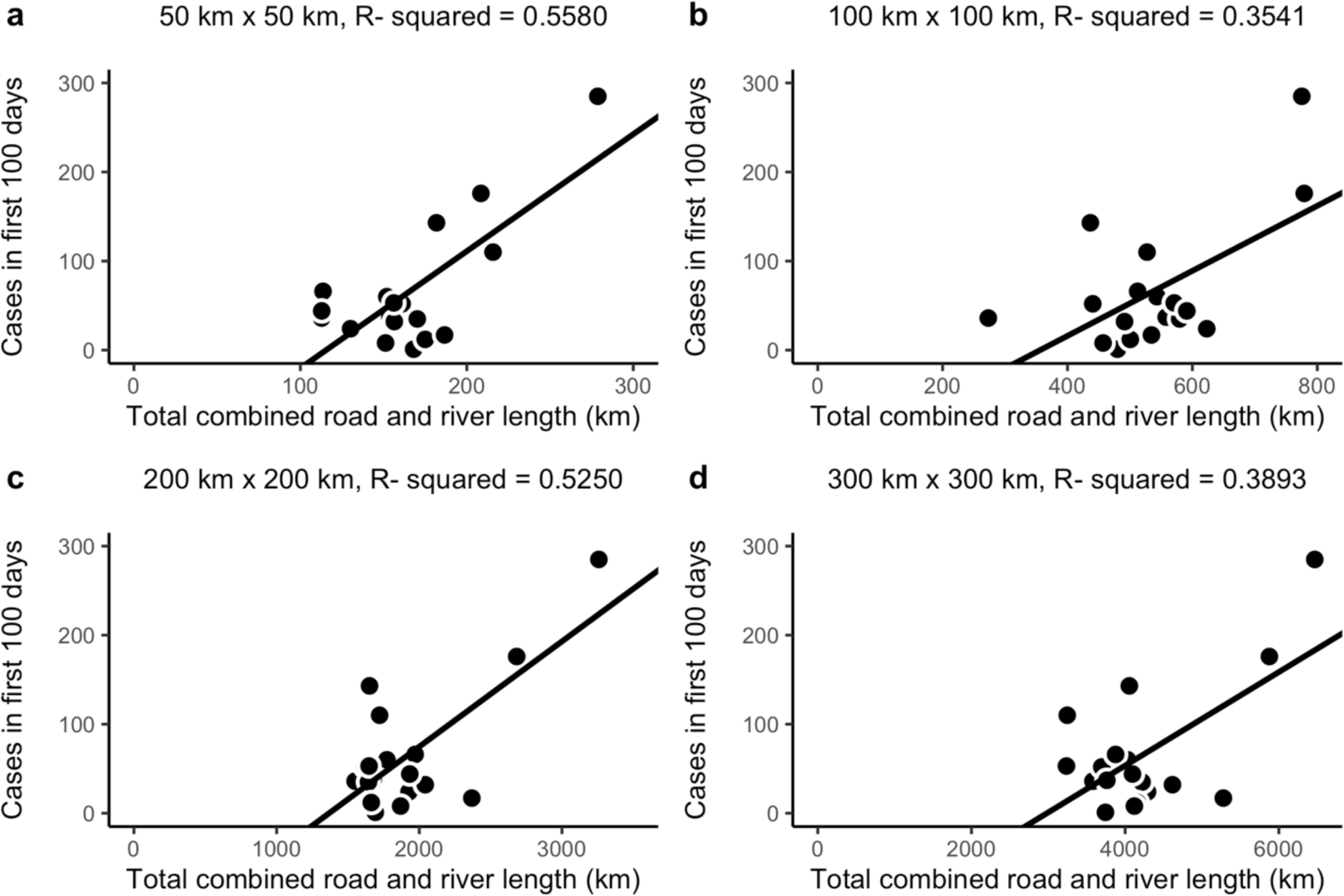
Linear Regression models of the total cases in the first 100 days and the total combined road and river length in km in a **(a)** 50 km x 50 km study area (R-squared = 0.5580, p = 0.0004), **(b)** 100 km x 100 km study area (R-squared = 0.3541, p = 0.0092), **(c)** 200 km x 200 km study area (R-squared = 0.5250, p = 0.0007), **(d)** 300 km x 300 km study area (R-squared = 0.3893, p = 0.0057).

## Notes

### Competing Interest Statement

The authors have declared no competing interest.

